# Preparation of robust Synthetic Control samples and their use in a metatranscriptomic clinical test

**DOI:** 10.1101/2023.04.28.23289273

**Authors:** Ryan Toma, Lan Hu, Guru Banavar, Momchilo Vuyisich

## Abstract

Metatranscriptomics (MT), or RNA sequencing, has the potential to revolutionize the field of molecular diagnostics. Due to the complexity of MT diagnostic models, positive and negative control materials for specific disease indications can be difficult to obtain. Controls must often be sourced directly from patients. This introduces logistical burdens, assay variability, and limits high throughput clinical laboratory operations. To overcome this limitation, we developed a method for generating Synthetic Control (SC) samples, which duplicate the nucleic acid signature of complex clinical specimens and produce the desired test outcome. SCs can be easily and cost-effectively produced in large quantities (>100,000 SCs per amplification cycle), enabling high throughput diagnostic testing. Here, we report the generation of Synthetic Positive Control (SPC) samples. SPCs were validated and implemented in a clinical laboratory. The SPCs produced robust positive signals (average OC risk score of 0.997) and high levels of reproducibility (%CV of 0.2%) in a high throughput automated CLIA laboratory. SCs are a novel and useful method for the generation of high quality controls for MT-based diagnostic tests, and their adoption could herald the widespread use of MT tests in molecular diagnostics.

## Introduction

Metatranscriptomics (MT), or RNA sequencing, is becoming an important analytical method in the field of screening and diagnostics, offering a comprehensive platform for the interrogation of a variety of chronic and communicable diseases ^1–4^. With its ability to generate large amounts of transcriptomic data, RNA sequencing has become an increasingly popular tool for identifying genetic mutations and variations^5^, detecting infectious agents^1^, understanding the role of human gene expression profiles in health and disease^6^, and elucidating the impact of the various human microbiomes in chronic diseases^7–9^. As MT technology continues to advance, its potential for improving the accuracy and speed of disease diagnosis is rapidly increasing, making it a potentially revolutionary tool for molecular diagnostics.

MT methodologies have already been used to develop diagnostic tests for a variety of chronic diseases that would not have been possible with prior methodologies. For example, MT has been used for accurate screening of oral and throat cancer ^3^, type 2 diabetes ^2^, irritable bowel syndrome ^10^, and autism spectrum disorder^4^. Despite the advantages of MT in disease diagnostics there are numerous challenges that must be overcome before these tests can be translated from research into the healthcare system.

Unlike other targeted diagnostic methods such as quantitative PCR or amplicon sequencing that are looking for a few gene mutations, most MT models utilize machine learning to identify hundreds of molecular features associated with an indication ^2,3,11^. While adequate positive or negative control materials can readily be generated for targeted methodologies by introducing the specific mutations or genes of interest, this is not feasible with the complex signals seen in MT disease models ^12–14^. Control material is an essential component of any test as it serves as a known reference point for the performance of the test and helps to ensure accuracy and reproducibility. However, obtaining large amounts of high-quality control material for clinical assays can be challenging, particularly for unbiased tests. The shortage of control material negatively affects the clinical validation of tests and the confidence in their results, and is a barrier for the widespread implementation of MT-based diagnostic tests.

Traditional control material for sequencing based assays is typically from patients that are known to be positive or negative for an indication of interest ^15,16^. However, the collection of these samples can be difficult and the sample is finite, which means that more control material from different patients must continually be obtained. This can result in significant logistical challenges. The need to continually obtain control material from different patients also raises questions about the stability, consistency, and reliability of the control source over time.

To address these challenges, our laboratory has developed Synthetic Controls (SCs) for MT assays. SCs can be made by unbiased amplification of total RNA extracted from a clinical specimen. This process allows for the generation of a highly controlled and standardized sample that mimics the RNA profile of the original sample, thus consistently returning a known test outcome. The use of SCs eliminates the need to continually obtain specimens from patients, as they provide an almost limitless source of synthetic control material.

Herein we present the methods for generating SCs and present data from the implementation of synthetic positive controls (SPC) for an oral and throat cancer screening test in a CLIA laboratory. The results demonstrate robust SPC performance that is clinically useful. We show that SPCs are a viable and effective approach to controlling MT tests and could be adopted as standard practice.

## Methods

### Ethics

This study (clinicaltrials.gov registration number NCT05451303) was conducted with a protocol and consent forms approved by an Institutional Review Board (IRB) accredited by the United States Health and Human Services. All participants consented to participate in the study.

### SC Preparation and Initial Validation

Two saliva samples from patients with clinically adjudicated oral cancer (OC) were used to generate SPCs. These samples were verified to test positive using CancerDetect, an oral and throat cancer screening test that is a clinically validated and licensed Laboratory-Developed Test (Viome Life Sciences)^3^. Total nucleic acids were purified, and DNA was degraded using DNase, followed by heat inactivation. Human and microbial ribosomal RNAs were removed using subtractive hybridization. The remaining transcripts were converted to cDNA with 5’ and 3’ PCR primer-annealing adapters appended (figure 1). To ensure that full length transcripts were recovered to mimic the RNA size distribution in the original sample, RNA fragmentation and cDNA size selection were not performed. The methods for SC generation up through the end of cDNA synthesis is identical to our laboratories previously published methods^17^. The cDNA was purified prior to PCR (AMPure XP Reagent, Beckman Coulter). The cDNA pool was amplified by PCR, where the forward primer contains a T7 promoter sequence while the reverse primer was designed to be as short as possible while maintaining a comparable melting temperature to the forward primer (figure 1). The resulting PCR product (F0) was used for *in vitro* transcription with AmpliScribe T7 High Yield Transcription Kit (Biosearch Technologies, AS3107) (figure 1). RNA from the transcription reaction was Dnase treated, cleaned up (AMPure XP Reagent, Beckman Coulter), and resuspended in nuclease free water.

**Figure 1:**
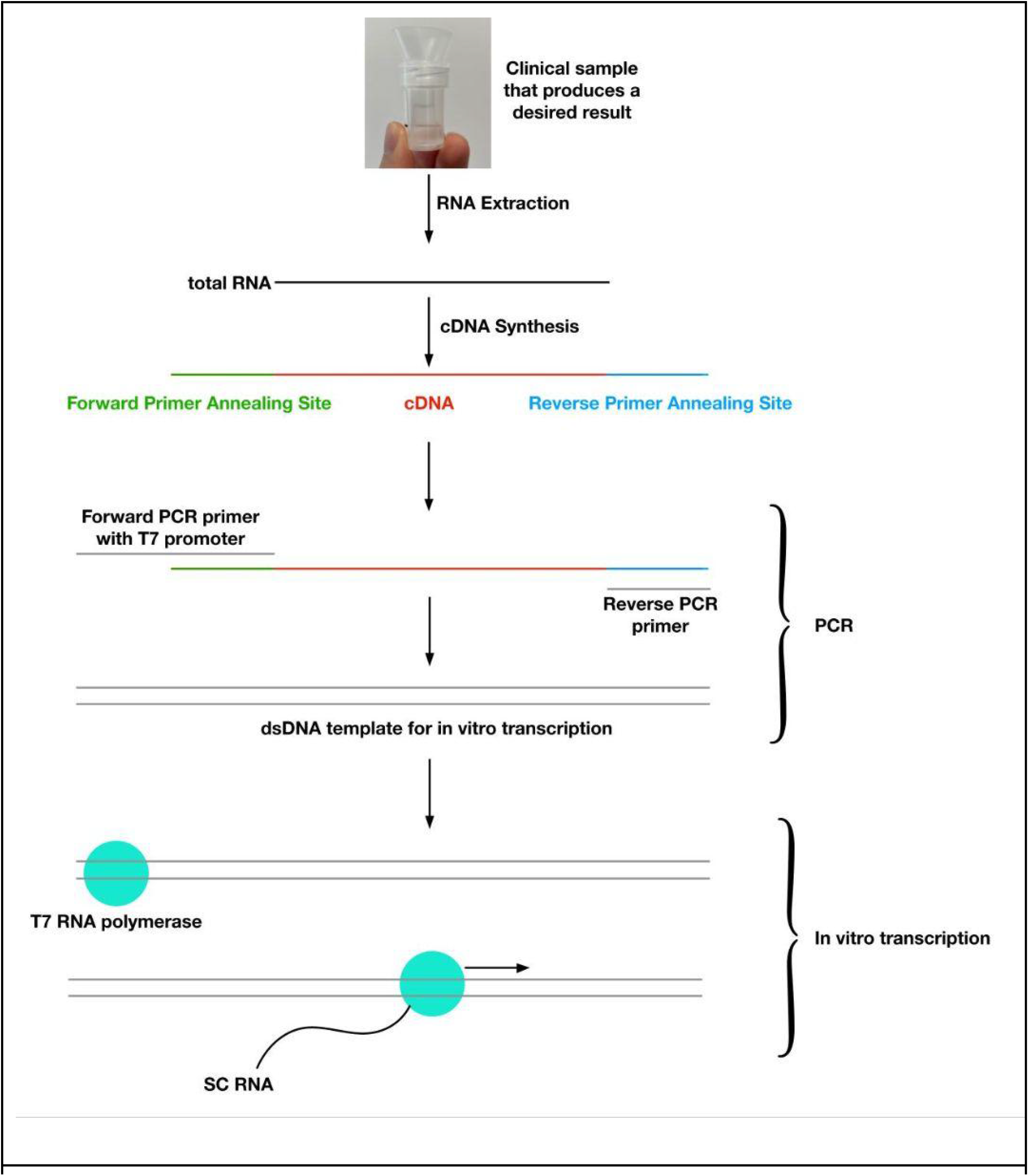
Preparation method for Synthetic Controls.

For the test of repeated amplification, the initial PCR product (F0) was purified, quantified, diluted, and re-amplified by PCR (figure 2). Purified dsDNA template (3 micrograms) was used as input for the next PCR reaction. This process was repeated three times (F1-F3). Each resultant F0-F3 dsDNA template was also *in vitro* transcribed as described above. Each SPC sample (150 ng of amplified RNA) was analyzed using the CancerDetect test in triplicate. In addition, 158 samples were collected from the general population and were analyzed through the CancerDetect test as a comparison cohort (all samples had ≥500,000 microbial ESD).

**Figure 2:**
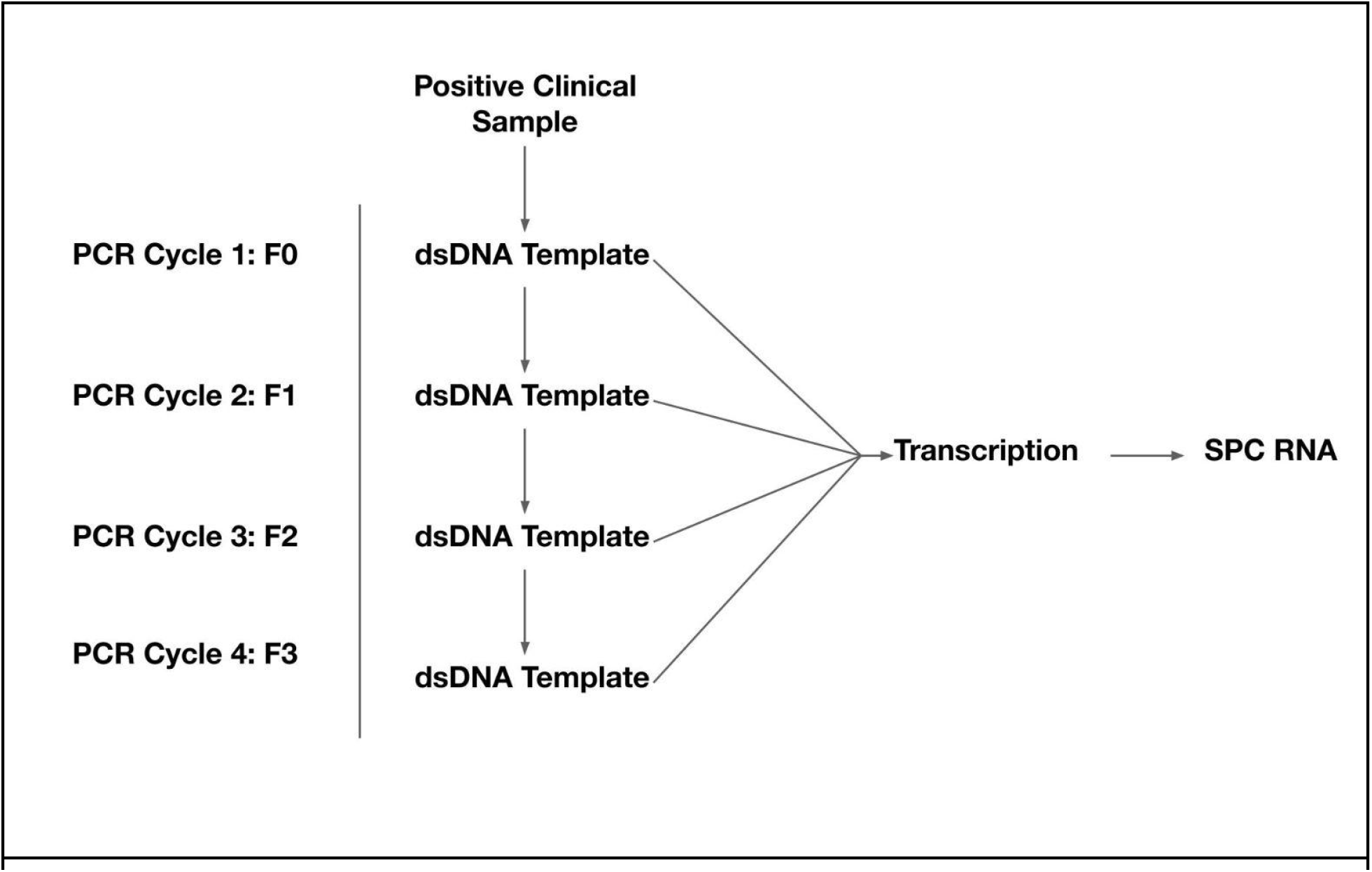
Diagram of repeated dsDNA template PCR amplifications.

### Library Preparation

SPC samples were analyzed using the CancerDetect test. The methods for this test have been previously published ^18^. Briefly, the method includes DNase treatment, non-informative RNA depletion, cDNA synthesis, size selection, and limited cycles of PCR for adding dual unique barcodes to each sample.

### SPC Clinical Validation

SPCs from F1 were generated for use in a clinical laboratory (Viome Life Sciences) using the methods outlined above. For the validation of SPCs, 18 replicates were processed through the CancerDetect test and their OC risk scores were calculated. The OC risk score is the predicted probability returned by the oral and throat cancer classifier, pre-trained and validated as previously described^3^. SPCs used in the clinical test had to have ≥10,000,000 total single reads, ≥500,000 microbial ESD, ≥1,000 KO richness, and ≥100 species richness to pass QC and be included in downstream analysis. All 18 replicates passed these QC criteria.

### Bioinformatics and Disease Classification

Our laboratory maintains a custom reference catalog which includes 32,599 genomes from NCBI RefSeq release 205 ‘complete genome’ category, 4,644 representative human gut genomes of UHGG^19^, ribosomal RNA (rRNA) gene sequences, and the human genome GRCh38^20^. These genomes cover archaea, bacteria, fungi, protozoa, phages, viruses, and the human host. The microbial genomes have 98,527,909 total annotated genes. Our laboratory adopts KEGG Orthology (KO) ^21^ to annotate the microbial gene functions using eggNOG-mapper^22^.

The microbiome pipeline maps paired-end reads to this catalog using Centrifuge^23^ for taxonomy classification (at any taxonomy rank). Reads mapped to the host genome and rRNA sequences are tracked for monitoring but excluded from further analysis. Reads mapped to microbial genomes are processed with an Expectation-Maximization (EM) algorithm^24^ to estimate the expression level (or activity) in the sample. Respective taxonomy ranks (strains, species, genus, etc.) can be easily aggregated from the genomes. For this study, we use species activity in the downstream analyses. These genome mapped reads are extracted and mapped to only gene or open reading frame (ORF) regions for molecular function or KO annotation and quantification.

We define the number of reads mapped to the microbiome as ‘microbial ESD’ (Effective Sequence Depth) to represent the usable portion of reads in a sample for microbiome. ESD only refers to the reads that have an identical match to the microbial database over contiguous 240 bps, which allows clinically validated, strain level taxonomic classification. Many additional reads align to the microbial ORF (open reading frame) database. The percentage of total reads aligning to our custom reference catalog is referred to as %Microbial ESD. OC risk scores were calculated based on our laboratories previously published machine learned (ML) classifier for oral and throat cancer that requires at least 500,000 microbial ESD to return a valid result^3^.

## Results

### SPC Metrics

Two patient samples were shown to be positive for Oral or Throat Cancer via the CancerDetect test and the diagnosis was confirmed by clinical examination. The predicted OC risk score from the CancerDetect test was 0.99 for donor SPC_1 and 0.89 for donor SPC_2. Each positive sample was then converted into a SPC.

The resulting dsDNA (F0) was used as a template for *in vitro* transcription or purified for repeated amplification and transcription (F1-F3). The average RNA and dsDNA yields of F0-F3 are reported in table 1.

**Table 1:** Average dsDNA template and RNA yields of F0-F3.

| **Amplification Cycle** | **dsDNA Template Yield (ng)** | **SPC RNA Yield per PCR Reaction (ng)** |
| --- | --- | --- |
| F0 | 453 | 119,590 |
| F1 | 839 | 181,163 |
| F2 | 546 | 311,276 |
| F3 | NA | 346,968 |

F0 generated sufficient dsDNA template for >150 additional PCR reactions, each of which can generate >100ug of SPC RNA. A single F0 PCR reaction is therefore capable of producing >15mg of F1 SPC RNA. The quantity of SPC RNA produced by the method is sufficient for >100,000 SPC samples, demonstrating the scalability of the method.

SPC samples generated ample sequencing data (table 2). %Microbial ESD was markedly suppressed with increased PCR cycling, indicating that repeated PCR amplifications reduces the amount of usable data. This can be easily overcome with increased sequencing depth.

**Table 2:** Average sequencing metrics for SPC samples.

| **Amplification**<br>**Cycle** | **Total Single Reads** | **Microbial ESD** | **%Microbial ESD** |
| --- | --- | --- | --- |
| F0 | 9,387,282 | 923,981 | 9.84 |
| F1 | 9,983,775 | 525,875 | 5.27 |
| F2 | 7,876,085 | 120,500 | 1.53 |
| F3 | 8,338,866 | 44,927 | 0.54 |

### SPC OC Risk Scores vs. Amplification Cycles

Even though the SPC_1 and SPC_2 OC risk scores were not significantly different between F0 and F1, F2, and F3 (p>0.05), there is a trend towards a reduction in the risk score with increased amplification cycles. The results indicate that SPCs should undergo a maximum of two additional rounds of PCR (up to F2) to minimize the degradation of the positive signal (figure 3A and 3B). However, this could be different for other tests. Even for this particular test, if the threshold for a positive test result is changed at some point in the future, re-amplification may or may not be appropriate.

**Figure 3:**
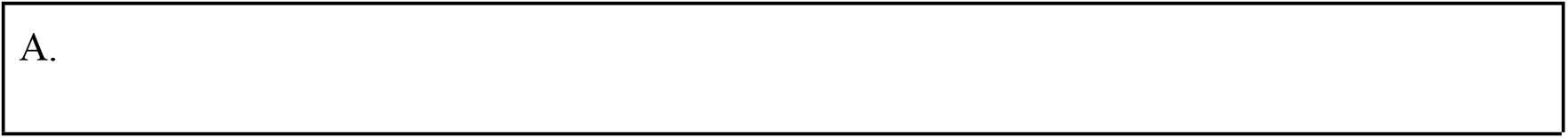
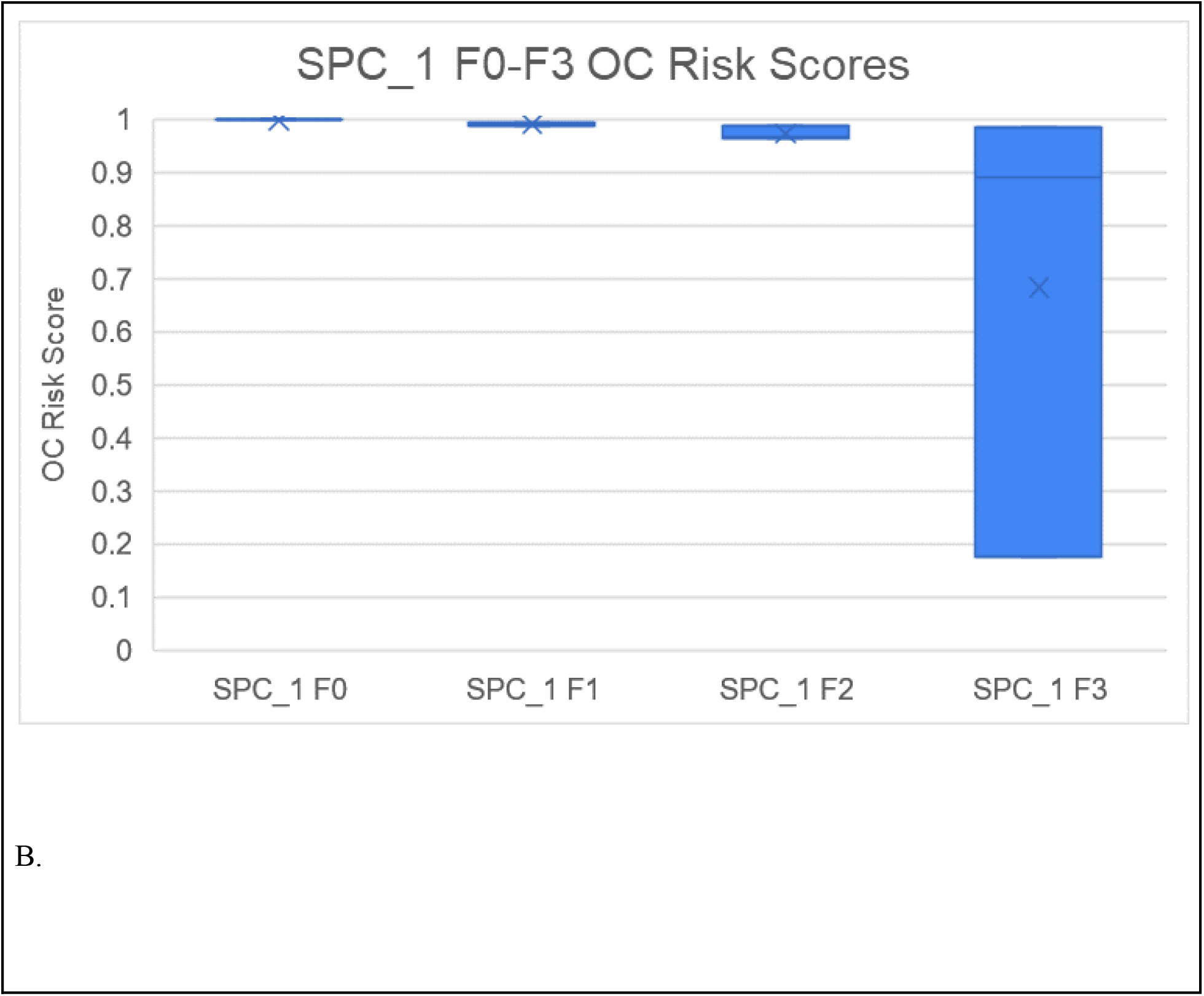
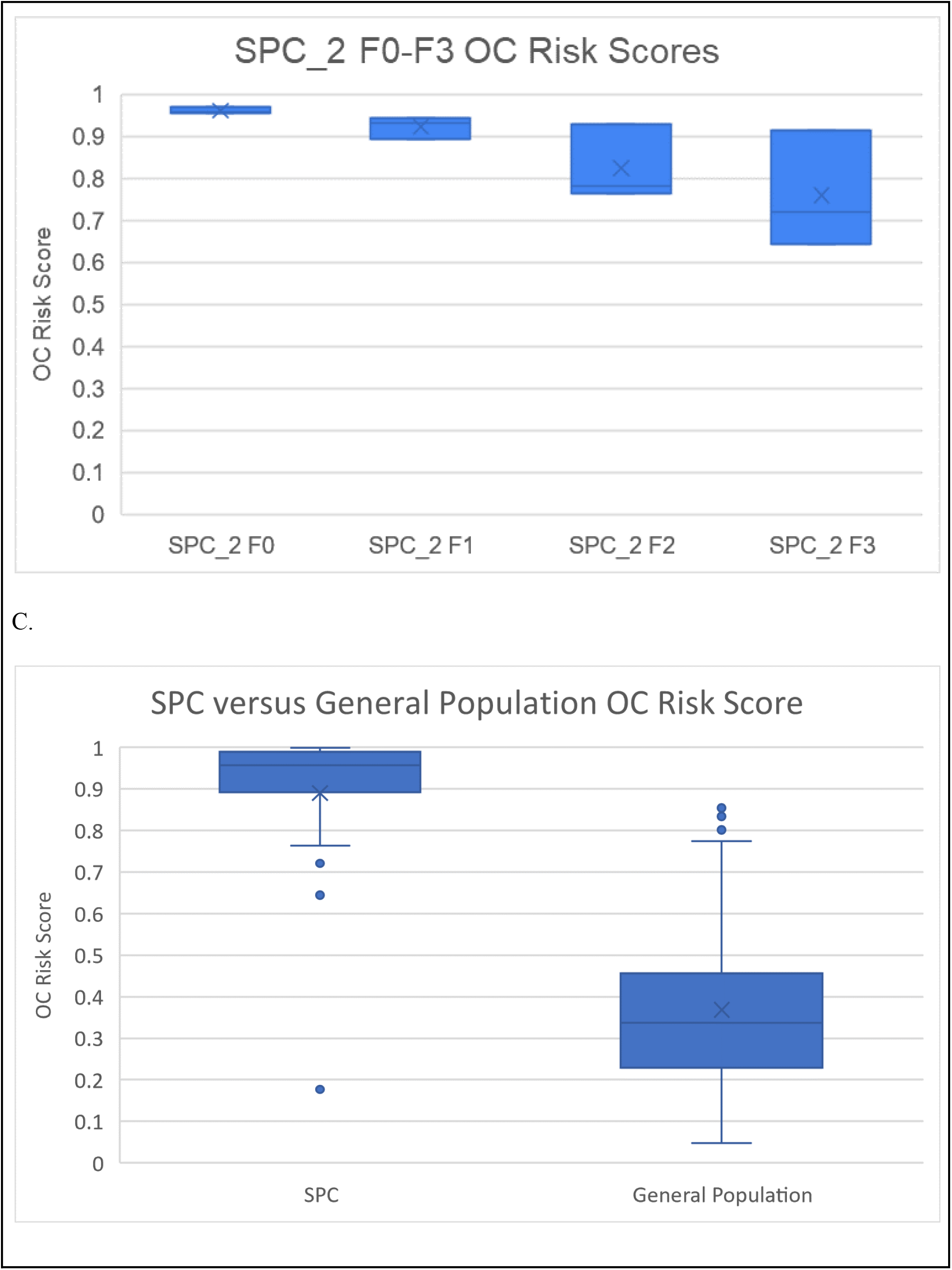
SPC performance is affected by repeated PCR amplification and produces robust positive signals.

All SPCs (SPC_1 F0-F3 and SPC_2 F0-F3, combined data) showed significantly increased positive signal compared to the general population (p<0.001, figure 3C). SPCs are effective at producing robust positive signals that are significantly higher than that of the general population even with repeated PCR amplification.

### SPC Implementation in a Clinical Test

SPCs were implemented in a clinical laboratory that performs a high throughput test for oral and throat cancer screening (CancerDetect, Viome Life Sciences). The average SPC OC risk score was 0.997 (standard deviation: 0.002) and the percent coefficient of variation between all replicates was 0.2%. These results demonstrate that SPCs can be utilized in a clinical test with high confidence in the SPC returning the desired outcome (positive).

## Discussion

The utilization of MT in diagnostic tests has the potential to significantly improve healthcare. However, the lack of suitable control materials remains a significant challenge in its clinical adoption. Currently, for MT diagnostic assays with complex molecular features, controls must be obtained from patients known to be positive or negative for an indication. However, this method is often problematic as sample collection can be difficult and the sample amounts are limited. This results in the need to continually obtain more control material from different patients, which can be logistically difficult.

In order to address these issues, our laboratory has developed a new method for producing control materials for MT assays. Synthetic Controls (SCs) can be created by amplifying the total RNA, which includes the diagnostic signatures, from samples that are known to be positive or negative. This allows for a near limitless source of control material that is easy and cost effective to produce, and eliminates the need for continuous collection of control material from patients.

One of the key advantages of SCs is their stability and robustness. Positive SCs (SPCs) were shown to produce positive signals even after multiple rounds of DNA template amplification (figure 3), indicating that the positive signal is stable and that the method does not introduce significant sources of bias. This makes SPCs ideal for use as control materials in MT diagnostic tests. In addition, SPCs are also cost-effective, as they eliminate the need for frequent collection of positive control material from patients and can be produced in large quantities in a controlled laboratory setting. Future research should seek to replicate these findings with negative SCs (SNCs).

We also demonstrate a clinical application of SPCs. SPCs were utilized as positive controls in a clinical test for an oral and throat cancer screening test (CancerDetect by Viome Life Sciences). The results demonstrate that SPCs can be translated into a clinical setting while maintaining their robust performance. Few research methods are ever implemented into clinical laboratories due to issues with high throughput scaling, reproducibility, and cost. SPCs overcome these challenges and have been shown to be a feasible, effective, and relevant control for an actual clinical MT test.

The concept of SC preparation can likely be applied to additional next generation sequencing (NGS) molecular biology diagnostic techniques such as metagenomics and amplicon sequencing. Metagenomics for example has already been used to generate models for a variety of conditions ^12,25–28^. These NGS assays would greatly benefit from access to improved control materials. Future research should seek to apply the methods in this paper to other fields of NGS diagnostics.

SCs offer a significant advancement in the field of MT diagnostics. The stability, robustness, and cost-effectiveness of SCs make them ideal for use as control materials in MT assays. By utilizing SCs, the field of molecular diagnostics can improve the accuracy and reliability of MT and potentially other NGS tests, thereby making them a mainstay in the modern healthcare system.

## Data Availability

All data produced in the present study are proprietary to Viome Life Sciences but may be available upon reasonable request to the authors

## Author Contributions

MV and RT designed and implemented the study. MV and RT developed the SC preparation method. RT performed the molecular analyses of the SPC samples. GB and LH performed data analysis. All authors contributed to data interpretation. All authors contributed to the writing of the manuscript.

